# The Psychological Impact of the COVID-19 Pandemic, A Meta-Analysis and General Population Study

**DOI:** 10.1101/2025.02.02.25321549

**Authors:** Derek C. Korff-Korn, Andrew Trauben, Elliot J. Rayfield

## Abstract

This study assessed the impact of the financial, social and health disruptions due to the pandemic on increased rates of depression, anxiety, obsessiveness and substance abuse tendencies. In the meta-analysis portion of the study, a systematic search was performed and resulted in twenty studies which met the appropriate relevance and quality. The degree of symptoms was assessed in comparison to certain variables such as sex, age and income. Given the diversity of countries in which these studies were sourced, a varying severity of pandemic and subsequent restrictions were observed which allowed for a more expansive comprehension of the correlation to mental health risks. The complementary aspect of the meta-analysis was a general population study performed in the later months of 2020, which surveyed New York City residents. The provided questionnaire included sociodemographic background, previous mental health issues as well as associated factors such as social isolation, financial burdens and health concerns. The investigated levels of depression, anxiety and traumatic stress exposed positive correlations between younger age and mental health symptoms, a greater impact on one’s mental health due to social isolation than health exposure, a significance of news exposure, a fluctuation in various issues of anxiety, depression and stress when assessing participants who had been infected, and a difference in time from change in lifestyle to expressed mental health symptoms.

## Introduction

With an indefinite pandemic a result of rising SARS-CoV2 infections globally, understanding characteristics of mental health risks, in the context of initial measures to undermine the spread of the virus as well as the direct effect of infections, remain vital.

## Methods

Methods and results were determined based upon the Preferred Reporting Items for Systematic Reviews (PRISMA) procedures (Moher).

### Study collection

Following PRISMA guidelines, a systematic search was performed on Pub Med Central, Google Scholar and Worldwide Science on October 17 2020. The eligibility criteria regarding search terms are as follows: (COVID-19 OR SARS-coV-2 OR severe acute respiratory syndrome coronavirus OR 2019nCoV OR HcoV-19) AND (Depression OR Anxiety OR Distress OR Substance Abuse) AND (Social OR Financial OR Health Concerns). The process of the systematic search and results are shown using the PRISMA flow diagram below (Fig. 1).

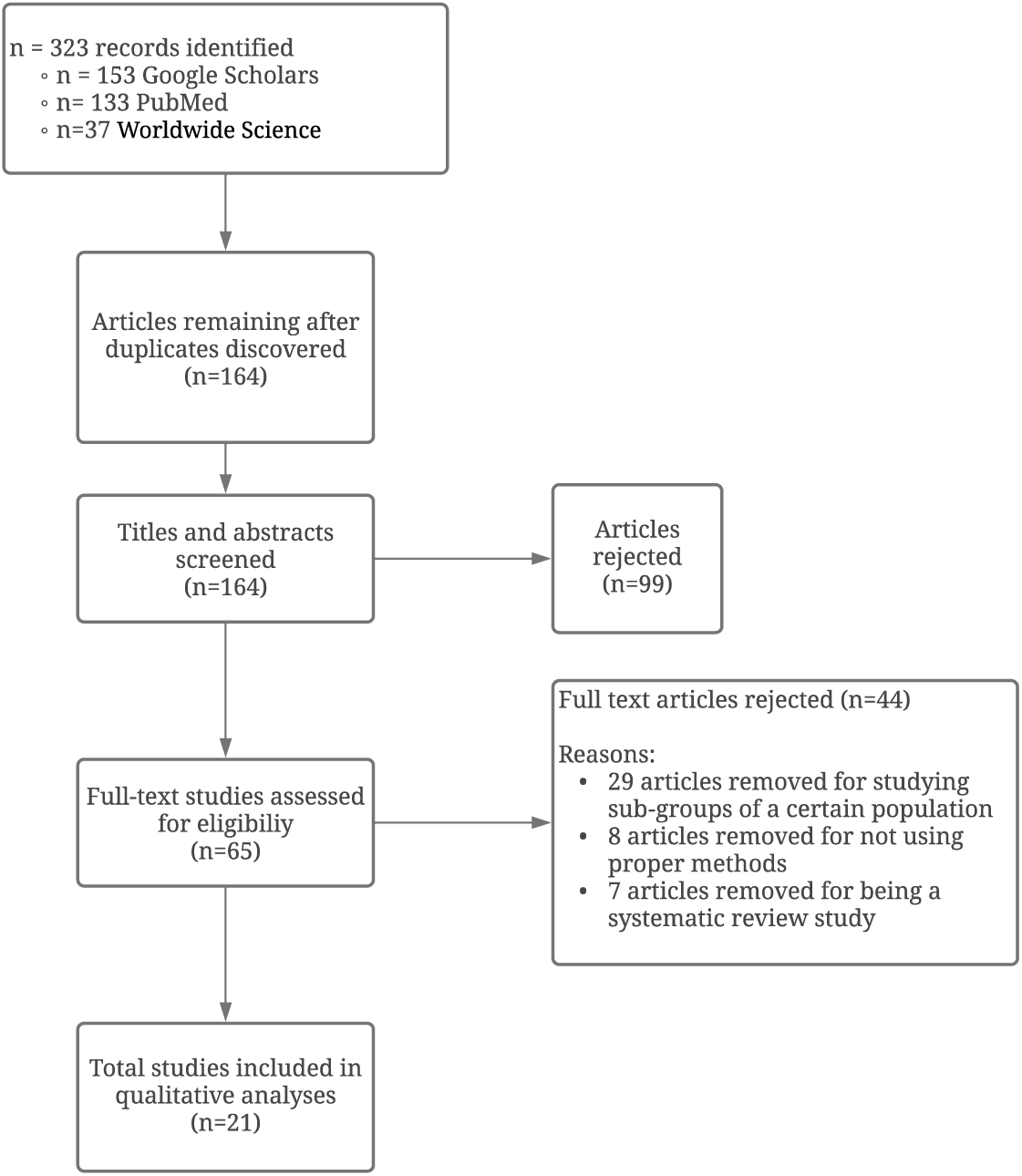

### Eligibility

After searching the detailed terms, the titles (and abstracts) were reviewed for relevance. Following the initial screening, full-text studies were assessed for the following criteria.

1. consistent with cross-sectional study, 2. regarded the mental health status of a population during the COVID-19 pandemic (January 2020-January 2021), 3. regarded the disruption to normal lifestyle (social, financial and health) caused by the pandemic as factors. Studies were rejected if 1. were not peer-reviewed, 2, exclusive to certain subgroups of populations such as students or frontline workers.

### Quality Control

In order to assess the quality of the selected studies the Newcastle-Ottawa Scale (NOS) was applied which asses the value of nonrandomized studies used in a systemic review; results of which are shown in Table 1a. The scale evaluates three areas to determine the significance of the sample: Outcome, Comparability and Selection. A total of seven categories are evaluated: representativeness of sample, sample size justification, comparability between respondents and non-respondents, ascertainments of exposure, comparability based on study design or analysis, assessment of the outcome, and appropriateness of statistical analysis. Each category had up to four questions attached to help determine value. A total of nine stars–3 in the Outcome section, 2 in the Comparability, and 4 in the Selection section–could be allotted.

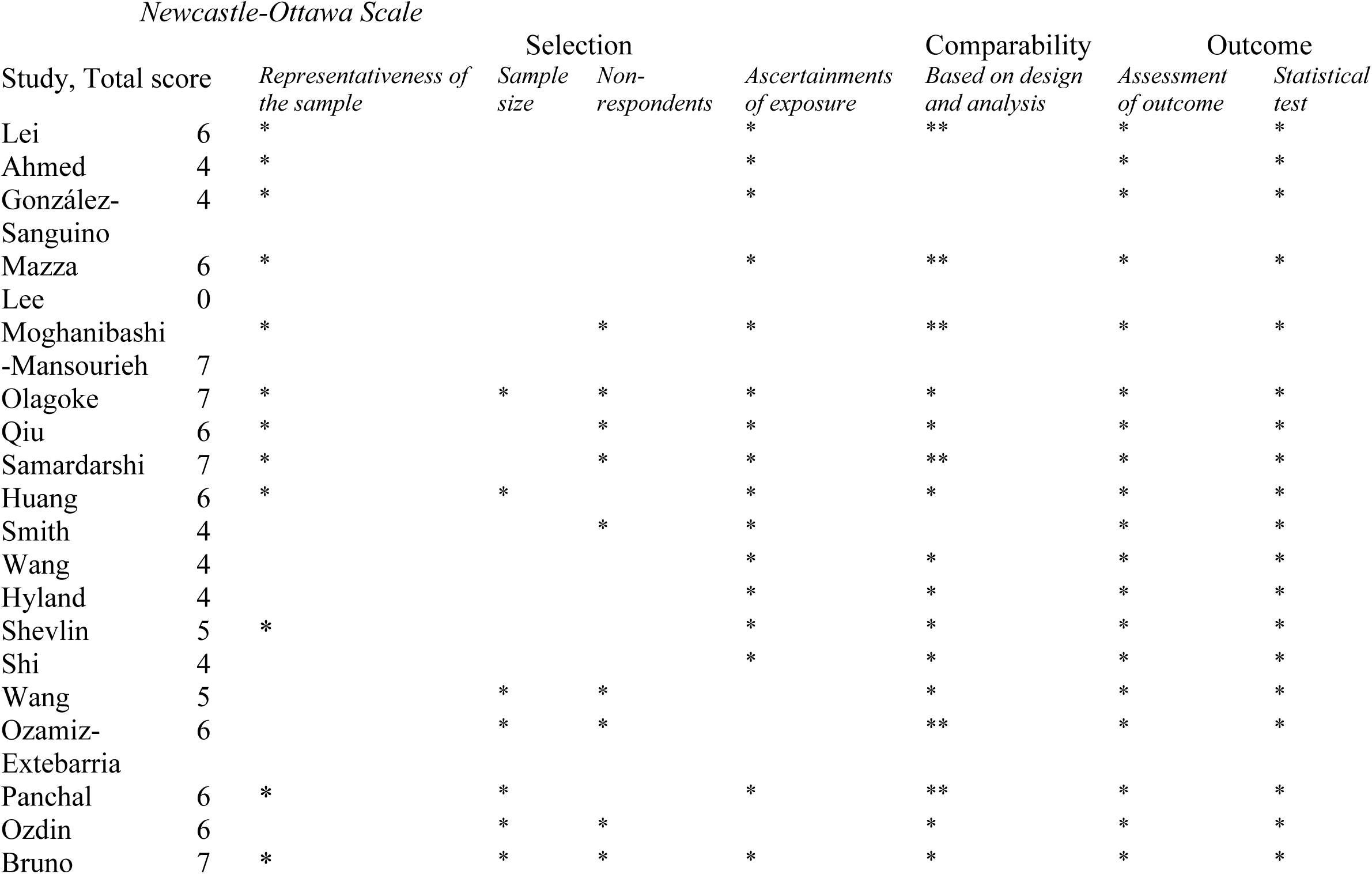

### Data discovery

To better organize and apply the selected studies, a form (Table 1a) was used to sort data. 1. First author, 2. Date (Month, Year), 3 Country of population, 3. Sample size, 4. Mean age 5. Assessment tools, 6. Prevalence of mental health symptoms (anxiety, depression, distress)

## Results

### Search

In total, 323 publications were discovered. After an initial screening, 164 articles were removed as duplicates. A further 99 articles were excluded after examining titles and abstracts. The remaining 65 articles were assessed: 29 studies were disqualified for including the review of a subgroup population, 9 articles for lacking proper study measures, and a final 7 articles for consisting of a meta-analysis. The full-text review considered 20 articles which met the eligibility standards.

### Study features

The primary details of the chosen articles are described in Table 1a. The sample size of 20 studies ranged from 318 to 56,932 participants with a total number of 155,919 and an average age of 36.8 years (17.4 SD). Female participants were a consistent majority throughout each study, representing 59.4% (n=92,546) of the population. The various studies were performed in nine different countries: China (n=7), Spain (n=2), United States (n=3), Italy (n=2), United Kingdom (n=2), Iran (n=1), Nepal (n=1), South Korea (n=1), Turkey (n=1), The studies consisted of the assessment of the mental impact of depression symptoms (n=7)– mostly due to social isolation and health concerns. A large majority (n=17) of the articles discussed the anxiety impact caused by the pandemic. However, 4 studies did not explicitly mention the total prevalence of mental health symptoms and only mentioned the frequency in comparison to associated risk factors.

**Table 1a.**
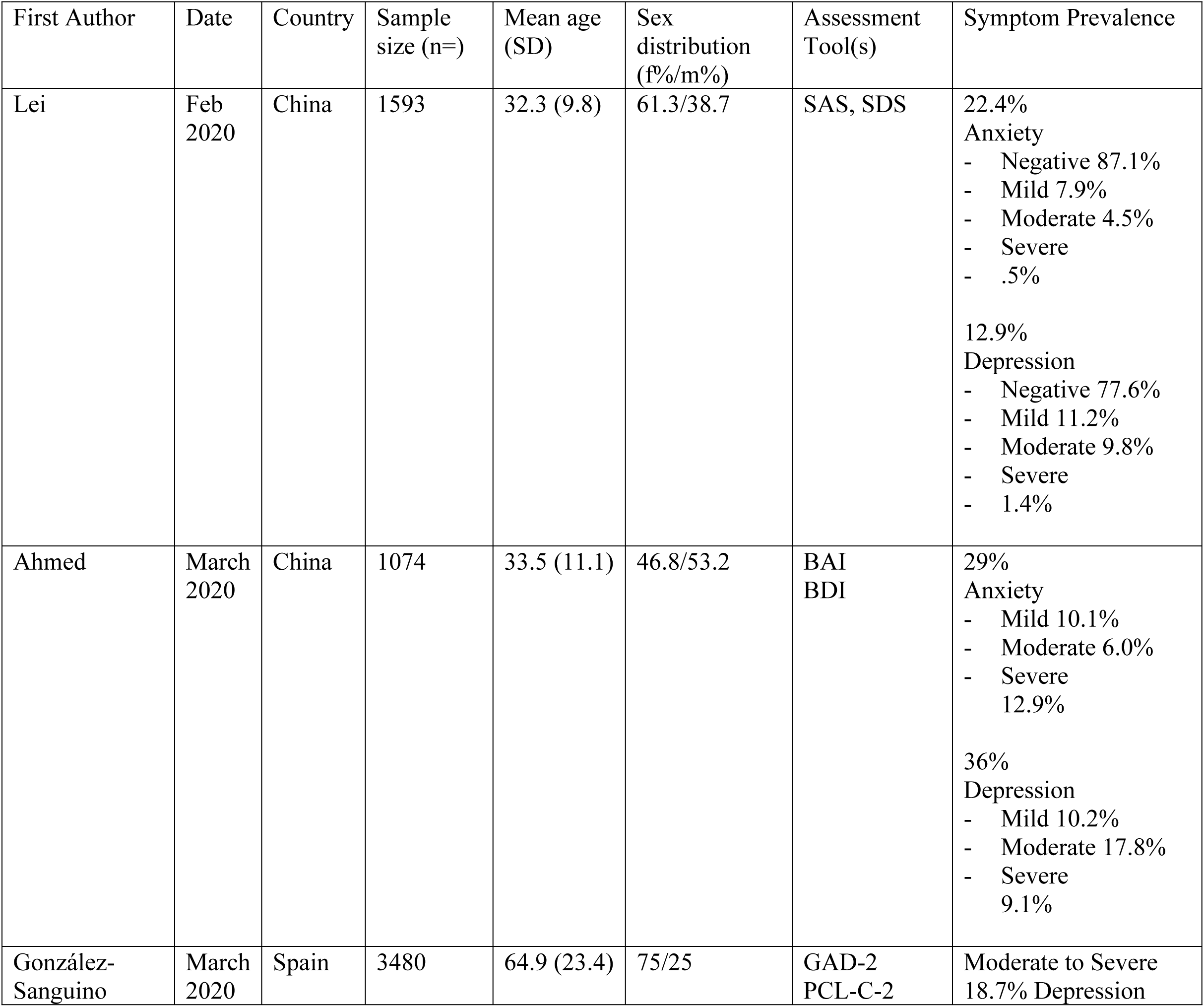
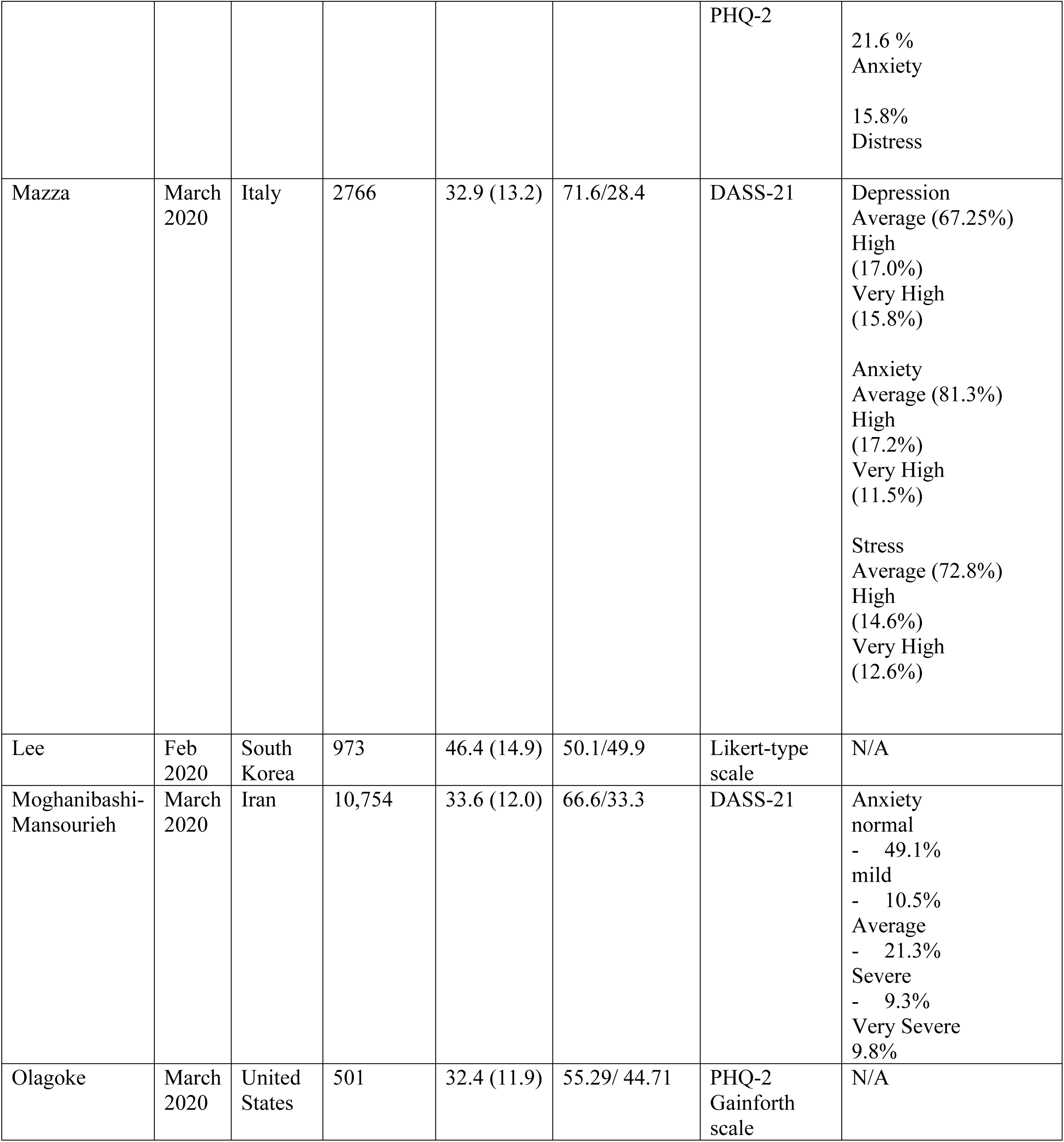
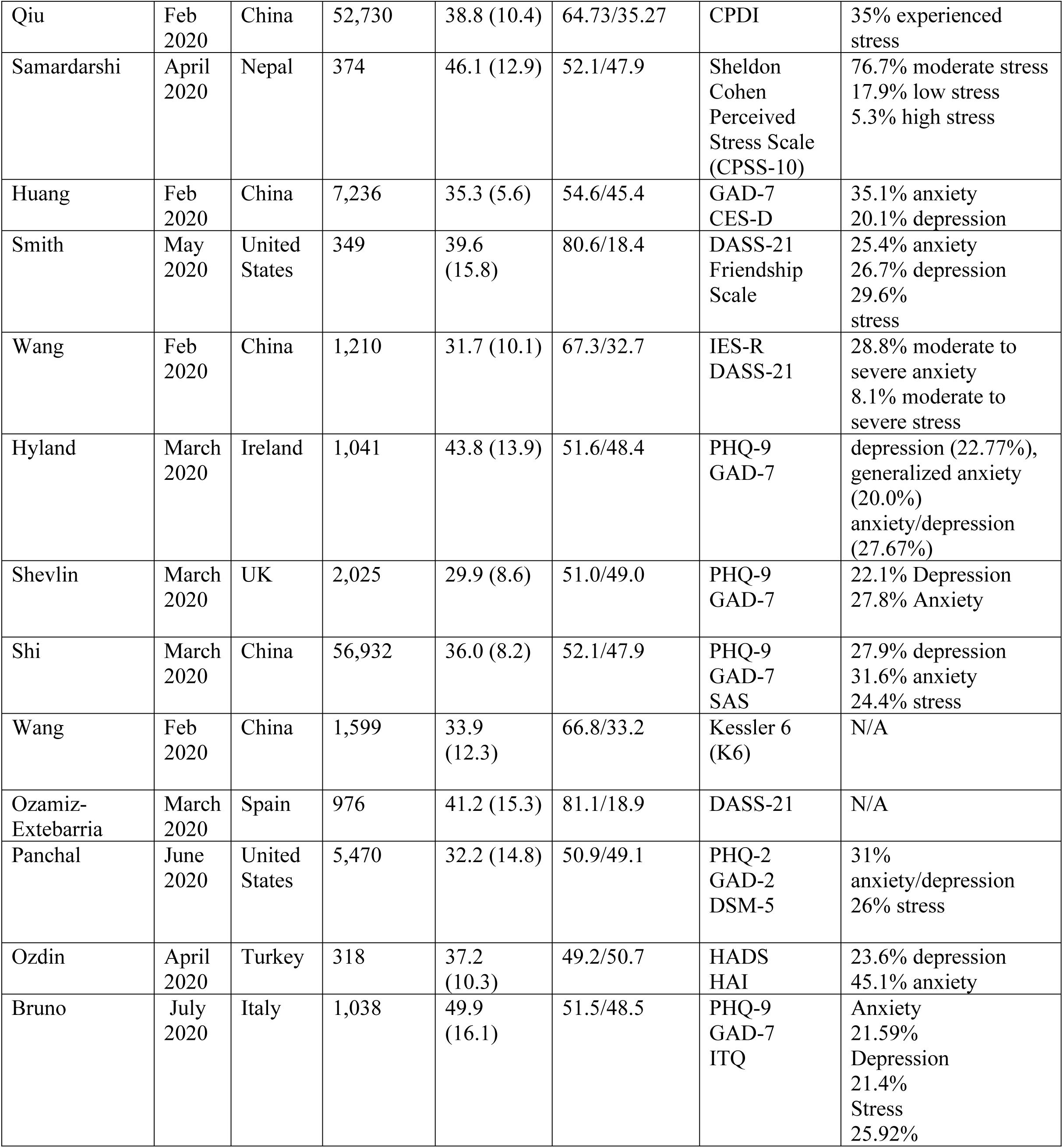

### Assessment Tools

A variety of scales were used to assess psychological symptoms. Anxiety was measured using the Generalized Anxiety Disorder 7-item (GAD-7), Self-rating Anxiety Scale (SAS) and the Health Anxiety Inventory (HAI). Depression was measured with the Patient Health Questionnaire (PHQ-9) and the Epidemiological Studies Depression scale (CESD). The Impact of Event Scale-Revised (IES-R), the Peritraumatic Distress Inventory (CPDI), the PTSD checklist (PCL-C-2) and the Kessler Psychological Distress Scale (K6) were used to determine the extent of distress in participants. The Depression, Anxiety and Stress Scale-21 (DASS-21) the Hospital Anxiety and Depression Scale (HADS) and the Diagnostic and Statistical Manual of Mental Disorders-5 (DSM-5) were also used to track multiple psychological disorders (CAMH, 2019).

#### Symptoms of depression and relevant factors^1^

Symptoms of depression were evaluated in 15 (out of 20) studies (Lei, 2020; Ahmed, 2020; González-Sanguino, 2020; Mazza, 2020; Huang, 2020; Smith, 2020; Wang, 2020; Hyland, 2020; Shevlin, 2020; Shi, 2020; Panchal, 2020; Ozdin, 2020; Bruno, 2020; González-Sanguino 2020; Olagoke, 2020). The prevalence of symptoms ranged from 12.9% to 36.0% (compared to a pre-pandemic frequency ranging from 3.6% to 7.2%). One must note that the self-report scales used in these studies are not definitive conclusions of an existing illness, such as clinical depression or any other, but rather a signifier of general depression symptoms.

Females were more at risk to developing depression symptoms in all 15 studies–ranging from an 11% to a 92.34% increase compared to their male counterparts. An increase in age (most consistently between age groups 18-30 vs. >50) suggested a decrease in symptoms, an extension of which is: students were found to also be more symptomatic as opposed to participants of other working status–such as employed or retired. Other factors which consistently suggested an increase in symptoms were having a lower annual income and being single (as opposed to married or in a relationship) cannot be regarded as products of the pandemic, but rather a pre-existing situation.

Each study was completed during a different version, experience of the pandemic– which include the accompanying circumstances: social, financial, and health–within the individual country (which is also influenced by the situation in the surrounding world). To assign significance to the prevalence of symptoms, the external factors–death rate, deaths, infections, lockdown procedures, government evaluation–were all considered for the time and place each of the 20 studies was conducted.

Throughout the various stages on the pandemic considered, from the beginning week of rising cases to the complete lockdown phase, the disruption of usual lifestyle, of which social interaction is most significant, seemed to be most impactful on depression symptoms. The first week of a lockdown, a stay-at-home order, observed in two articles (Ozamiz-Extebarria, 2020; Hyland, 2020), was enacted in response to alarming COVID-19 cases: in Spain all 50 provinces had confirmed cases, with a peaked death rate; in Ireland and the surrounding UK, a peak of 6,000 daily cases had been reached). The studies revealed increased depression due to social isolation, or at least lack of social deprivation a protective measure, such as the normal school environment replaced with an online format. The symptoms within distinct age groups also suggests the disruption in life is more prominent in young adults; depression symptoms between participants 18-25 years vs. >61 years was 22.7% vs. 6.4%, a 238% increase (Ozamiz-Extebarria, 2020). Older participants are less impacted by the lockdown order, as they are more likely to be retired and not as immersed in social activity as so social disruption was not as apparent. The rising cases and deaths did not seem to be related to depression symptoms.

The most drastic comparison is that of participants who were previously infected: 56.3% of said participants experienced depression symptoms compared to the 27.1% prevalence in symptoms for those who had not been infected experienced a (Shevlin, 2020).

However, studies which tracked participants in later stages of the pandemic in which most non-essential activities had been suspended for months, if not weeks, depression symptoms were directly corelated with health topics: confirmed/suspected with COVID-19, Family/friends infected, and a pre-existing condition (Shi, 2020; Shevlin, 2020). The general population described an increased in depression prevalence by 4.8% due to quarantine. Yet within those participants who went to work experienced 1.8% less symptoms than those who worked from home. Specifically, frontline workers only experienced an increase of symptoms of 11% by people working from home (Shi, 2020).

Social isolation contributed to an increased frequency of depression. When exposed to COVID-19 the depression (moderate to severe) prevalence was 18.4%. 9.0% of participants who reported moderate stress symptoms and were exposed to the general population experienced depression. 9.8% of people experienced depression when under quarantine. (Shi, 2020)

Regarding financial issues, depression symptoms were more than double in frequency when household income was less than $15,000 as opposed to >$60,000. When discussing the direct impact of the pandemic, lost income increased symptoms from 23.6% to 36.9% (Shevlin, 2020).

#### Symptoms of anxiety and relevant factors^1^

Anxiety symptoms were considered in 18 out of the 20 studies, with a prevalence of moderate to severe anxiety symptoms ranging from 10.2% to 45.1% (Lei, 2020; Ahmed, 2020; González-Sanguino, 2020; Mazza, 2020; Lee, 2020; Moghanibashi-Mansourieh, 2020; Olagoke, 2020; Huang, 2020; Smith, 2020; Wang, 2020; Hyland, 2020; Shevlin, 2020; Shi, 2020; Wang, 2020; Ozamiz-Extebarria, 2020; Panchal, 2020; Ozdin, 2020; Bruno, 2020). Symptoms of anxiety are frequently accompanied with symptoms of depression and in 2 studies, the prevalence of symptoms associated with both illnesses were combined (Panchal, 2020; Hyland, 2020): 31.0% and 27.7% respectively.

13 out of the 14 studies which detailed the sex distribution of anxiety prevalence indicated a correlation with female participants, ranging from a 12.1% to 32.3 increased compared to male participants.

Anxiety symptoms were associated with the following health factors: Taking cautious measures such as wearing a mask outside and not touching public objects (regarding the DASS-21, out of 20 points, the respective correlations with such behaviors were -0.43 and -0.53 (Wang, 2020)) were all protective in limiting symptoms; as well as an increased self-perceived health and lack of pre-existing conditions (Bruno, 2020; Ozdin, 2020; Olagoke, 2020; Mazza, 2020; Lei, 2020). Living near urban as opposed to rural areas, or highly infected provinces was a factor in increased anxiety. For instance, participants living in Hubai experienced anxiety symptoms twice as often as those in the surrounding provinces (Ahmed, 2020; Ozdin, 2020).

Prevalence increased if a family member or friend contracted the illness regardless of the participants proximity to the infected. The most drastic comparison is that of participants who were previously infected: 12.5% experienced anxiety symptoms, and 21.5% of those who had not been infected experienced symptoms of anxiety (Shevlin, 2020).

Financially, participants of lower income ($15,000 as opposed to >$60,000) reported an increase in symptoms from 22.9% from 18.8%. Regarding direct consequences of the pandemic, lost income was associated with 23.5% as opposed to 20.0% from participants who did not experience a financial loss (Shevlin, 2020).

Decreased age was also a factor in prevalence of symptoms. (Hyland, 2020; Huang, 2020; González-Sanguino, 2020; Lei, 2020). In one study, participants experienced a -0.23pt correlation in the GAD-7 (21 points) to years in age (Bruno, 2020). Anxiety had a 37% higher prevalence in participants 18-25 years old than those older than 50 (Ozdin, 2020). Anxiety was 96.2% higher in participants 18-25 years old than those older than 60 (Shevlin, 2020).

Anxiety regarding health concerns was promoted by the consistent stream of information through social media and news. There was a clear correlation between an increased level of focus on the pandemic to increased prevalence of anxiety; a nearly 200% uptick in symptoms was observed when participants consistently followed the pandemic on the news as opposed to never (Moghanibashi-Mansourieh, 2020; Huang, 2020).

Conversely, participants who were properly informed and had positive self-perceived knowledge regarding the pandemic were less likely to experience symptoms than those who were over-informed (Lei, 2020). In one study, the correlation of being properly informed to anxiety symptoms (using the GAD-2, 21 points) was -0.233, as opposed to being overinformed– a correlation of 0.15 (Ahmed, 2020).

#### Symptoms of psychological stress/distress and relevant factors^1^

Regarding distress/stress symptoms, prevalence ranged from 15.8% to 36.1%. 9 of the 20 studies studied the traumatic effect of the pandemic; a majority of which discovered a prevalence above 25% (Shi, 2020; Ozamiz-Extebarria, 2020; Panchal, 2020; Bruno, 2020; Qiu, 2020; Samardarshi, 2020; Smith, 2020; Mazza, 2020). Compared to the measurement of other psychological symptoms (anxiety and depression), there was a more diverse use of scales including: ASDS, DSM-5, PCL-C-2, ITQ, CPSS-10, CPDI and the K6. There was no discernable correlation between the female and male population of the 6 studies which detailed a sex distribution. In one study females were 23% less likely to experience stress, however in another the female population was 31.8% more likely to experience stress^1^ (Shevlin, 2020; Bruno, 2020).

Leaving home to work increased stress: there was a positive correlation of 1.55 on the DASS-21 point scale (Mazza, 2020); Migrant workers, who traveled by public transportation, experienced the highest score on the CPDI scale, with a mean of 31.89 relative to the general population average of 23.6 (Qiu, 2020); and a prevalence ratio of 1.52 comparing employees who left home to work and stay at home workers (Panchal, 2020).

Health concerns consistently were associated with higher stress: A participant who had a pre-existing condition experienced 30.9% more stress than participants who did not (Bruno, 2020). A family member/friend infected with COVID-19 increased stress by 120%. Any proximity to COVID-19 infected patients increased stress by 13.5% (Shi, 2020).

There is a small inconsistency among the age distribution. One study discovered significantly lower stress among the younger population (18-40) than participants older than 50 years (Qiu, 2020). People under 18 years had the lowest CPDI scores (a mean of 14.83 compared to the general population mean of 23.6 and the 27.8 mean for participants above 60 years). The majority of the 5 remaining studies which revealed an age distribution revealed a consistent higher stress level in the younger population relative to the older population ranging from a 19% to a 267.6% increase. (Ahmed, 2020; Samardarshi, 2020; Smith, 2020; Shi, 2020; González-Sanguino, 2020; Wang, 2020).

Participants who were less concerned about the media reports regarding the pandemic experienced 23.1% of the stress experienced by participants who reported they were extremely concerned with the news and social media reports (Wang, 2020).

Despite omnipresent health concerns, social isolation contributed to more a frequent stress prevalence. When exposed to COVID-19 the stress (moderate to severe) prevalence was 39.6%. 20.8% stress participants who were exposed to the general population experienced stress. 23.9% of people experienced stress when under quarantine. (Shi, 2020) Regarding specifically loneliness, there was a 29% increase of stress for participants who experienced social isolation, which was determined using the UCLA Loneliness Scale (Ozamiz-Extebarria, 2020).

There was a slight correlation between lost income and stress. The stress observed for participants with an annual income of $15,000 was 12.9% and for those of more than $60,000, average symptoms was 12.0%. 22.2% of participants who lost wages experienced stress compared to the 14.2% of participants who did not lose any income.

33.3% of participants previously infected with COVID-19 experienced traumatic stress compared to the 16.4% of participants not infected who experienced symptoms. (Shevlin, 2020). Although health risks were noticed to raise stress levels, avoiding such risks and remaining in quarantine was more detrimental to the psychological health than being more exposed.

## General Population Study

### Methods

The research was performed between November 7 2020 and December 20 2020 with a population consisting of individuals living in New York City, New York. The study was distributed through various social media platforms (Instagram, WhatsApp and Facebook) and completed using an online questionnaire. The sample size (n=96) was determined given the number of responses. The 45-item questionnaire included sociodemographic background, previous mental health issues (anxiety, depression, distress and substance abuse) as well as associated factors such as social isolation, financial burdens and health concerns. The scales included to determine severity and frequency of symptoms were the self-report scale for the Generalized Anxiety Disorder-7 (GAD-7), the Patient Health Questionnaire-9 (PHQ-9), the Perceived Stress Scale (PSS) and the Drug Addiction Assessment (DAA).

GAD-7 is a self–assessment scale developed by Dr. Robert Spitzer, which is used to determine anxiety levels. It consists of 7 questions, each of which is scored from 0-3 (not at all-nearly every day). The lowest score is a 0 and the highest score is a 21. The scale’s scoring was altered for a 0-4 range from 0-40.

PHQ-9 is a self-assessment scale developed by Dr. Kurt Kroenke, which is used to determine depression levels. It consists of 9 questions, each of which is scored from 0-3 (not at all-nearly every day). The lowest score is a 0 and the highest score is a 27. The scale’s scoring was altered for a 0-4 range from 0-28.

PSS is a self-assessment scale developed by Sheldon Cohen, which is used to determine depression levels. It consists of 10 questions, each of which is scored from 0-4 (not at all-nearly every day). The lowest score is a 0 and the highest score is a 40.

DAA is a self-assessment scale used to determine symptoms of addiction. It consists of 9 questions, each of which is scored from 0-4 (not at all-nearly every day). The lowest score is a 0 and the highest score is a 36.

Social isolation, financial burdens and health concerns were evaluated through a set of questions which was set on a final scale ranging from 0-10. For social isolation such range means: no impact to complete isolation. For financial burdens such range means: no impact to insolvent. For health concerns such range means: no concern to constant concern of risk of death (CAMH Foundation, 2019).

Results were expressed as mean (standard deviation) or N (%). Data was grouped into variables including gender, age and occupational status. In some tables, participants were grouped as individuals with a specific anxiety, depression, stress or addiction symptom based on cut-off scores; in this case the symptom was only regarded if it was considered moderate to severe–experienced “more than half the days” or “nearly every day”. Multiple linear regression was applied to identify relationships between the factors discussed and reported mental health symptoms.

### Results

A total of 96 individuals above the age of 18 completed the questionnaire online, all of whom lived in an urban area. 64 (66.6%) of the population is female and the mean age is 37.6 (STD ± 19.1 years). 45 (46.8%) participants are students, 62 (64.6%) are employed and 7 (7.3%) are retired (Table 1b). 31 (32.2%) participants met the cut of score for anxiety symptoms, 29 (30.2%) for depression and 31 (32.2%) for stress. Participants’ mean GAD-7, PHQ-9 and PSS score were 14.1 (STD ± 9.7), 11.4 (STD ± 7.3) and 14.2 (STD ± 12.2), respectively. Of the three associated factors observed, social isolation was most rampant with an average score of 7.8 (STD ± 4.9), financial burdens 2.0 (STD ± 1.6) and health concerns 7.0 (STD ± 3.2).

**Table 1b.**
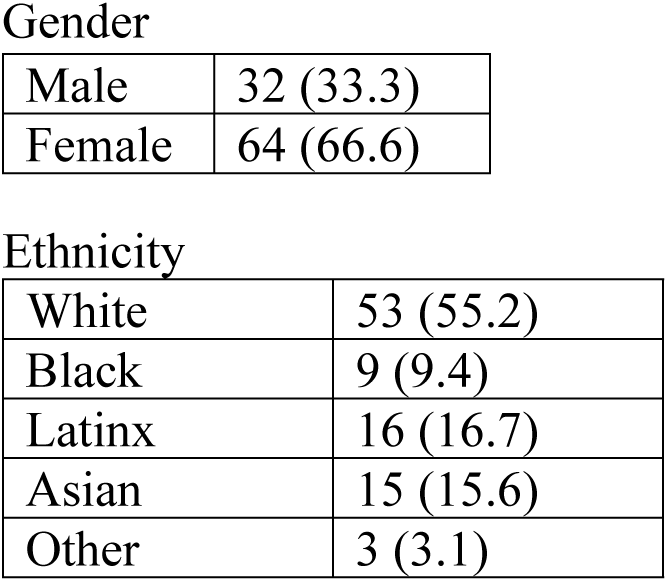
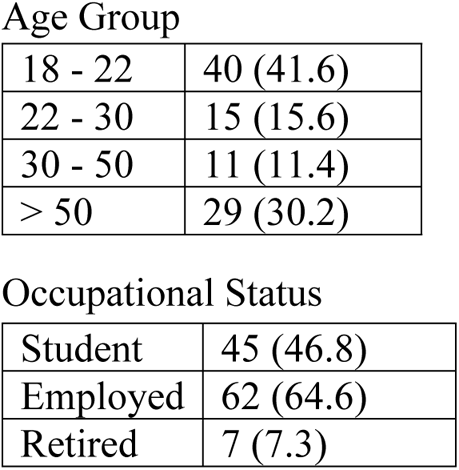
Sociodemographic and clinical features of the participants N (%)

Gender
|  |  |
| --- | --- |
| Male | 32 (33.3) |
| Female | 64 (66.6) |
| Ethnicity |  |
| White | 53 (55.2) |
| Black | 9 (9.4) |
| Latinx | 16 (16.7) |
| Asian | 15 (15.6) |
| Other | 3 (3.1) |
| Age Group |  |
| 18 - 22 | 40 (41.6) |
| 22 - 30 | 15 (15.6) |
| 30 - 50 | 11 (11.4) |
| > 50 | 29 (30.2) |
| Occupational Status |  |
| Student | 45 (46.8) |
| Employed | 62 (64.6) |
| Retired | 7 (7.3) |

**Table 2b.** Comparisons of participants in terms of PHQ-9, GAD-7 and PSS.

|  | Anxiety |  |  |  | Depression |  |  |  | Stress |  |  |  |
| --- | --- | --- | --- | --- | --- | --- | --- | --- | --- | --- | --- | --- |
|  | Non-Significant (0-10) | Mild (11-20) | Moderate (21-30) | Severe (31-40) | Non-Significant (0-7) | Mild (8-14) | Moderate (15-21) | Severe (22-28) | Non-Significant (0-10) | Mild (11-20) | Moderate (21-30) | Severe (31-40) |
| Pre-Pandemic | 29 (30.2) | 30 (31.25) | 32 (33.3) | 5 (5.2) | 36 (37.5) | 28 (29.2) | 23 (24.0) | 8 (8.3) | 9 (9.4) | 22 (22.9) | 52 (54.2) | 11 (11.5) |
| During Pandemic | 28 (29.2) | 27 (21.1) | 19 (19.8) | 12 (12.5) | 38 (39.6) | 31 (32.3) | 19 (32.3) | 12 (12.5) | 37 (38.5) | 28 (29.2) | 18 (18.8) | 13 (13.5) |

Gender
|  |  | Anxiety |  |  |  |  | Depression |  |  |  |  | Stress |  |  |  |  |
| --- | --- | --- | --- | --- | --- | --- | --- | --- | --- | --- | --- | --- | --- | --- | --- | --- |
|  |  | Non-Significant (0-10) | Mild (11-20) | Moderate (21-30) | Severe (31-40) | Mean (SD) | Non-Significant (0-7) | Mild (8-14) | Moderate (15-21) | Severe (22-28) | Mean (SD) | Non-Significant (0-10) | Mild (11-20) | Moderate (21-30) | Severe (31-40) | Mean (SD) |
| Male | 32 (33.3) | 14 (43.8) | 8 (25.0) | 6 (18.75) | 4 (12.5) | 16.8 (12.8) | 14 (43.8) | 11 (34.4) | 4 (12.5) | 3 (9.4) | 19.46 (8.34) | 15 (36.8) | 8 (25.0) | 5 (15.6) | 4 (12.5) | 27.31 (22.6) |
| Female | 64 (66.6) | 24 (35.9) | 19 (29.7) | 13 (21.9) | 8 (12.5) | 20.4 (18.1) | 20 (31.3) | 20 (31.3) | 15 (23.4) | 9 (14.1) | 23.84 (13.5) | 22 (34.4) | 20 (31.3) | 13 (20.3) | 9 (14.1) | 30.48 (24.0) |

Age group
|  |  | Anxiety |  |  |  |  | Depression |  |  |  |  | Stress |  |  |  |  |
| --- | --- | --- | --- | --- | --- | --- | --- | --- | --- | --- | --- | --- | --- | --- | --- | --- |
|  |  | Non-Significant (0-10) | Mild (11-20) | Moderate (21-30) | Severe (31-40) | Mean (SD) | Non-Significant (0-7) | Mild (8-14) | Moderate (15-21) | Severe (22-28) | Mean (SD) | Non-Significant (0-10) | Mild (11-20) | Moderate (21-30) | Severe (31-40) | Mean (SD) |
| 18 - 22 | 40 (41.6) | 11 (27.5) | 14 (35) | 9 (22.5) | 6 (15.0) | 27.32 (18.5) | 14 (35.0) | 10 (25.0) | 10 (25.0) | 6 (15.0) | 20.4 (15.1) | 11 (27.5) | 17 (37.8) | 6 (15.0) | 6 (15.0) | 22.6 (16.8) |
| 22 - 30 | 15 (15.6) | 6 (40.0) | 5 (33.3) | 2 (13.3) | 2 (13.3) | 21.0 (16.0) | 4 (26.7) | 6 (40.0) | 2 (13.3) | 3 (20) | 15.3 (12.8) | 6 (40.0) | 2 (13.3) | 5 (33.3) | 2 (13.3) | 19.9 (15.4) |
| 30 - 50 | 12 (12.5) | 6 (50) | 3 (25) | 2 (16.7) | 1 (8.3) | 16.82 (12.4) | 5 (41.7) | 4 (36.4) | 2 (18.2) | 1 (9.1) | 11.7 (5.1) | 6 (50) | 2 (16.7) | 2 (16.7) | 2 (16.7) | 18.0 (14.1) |
| > 50 | 29 (30.2) | 15 (51.7) | 5 (17.2) | 6 (20.7) | 3 (10.3) | 12.76 (10.8) | 11 (37.9) | 11 (37.9) | 5 (17.2) | 2 (6.9) | 7.1 (6.0) | 14 (48.3) | 7 (24.1) | 3 (10.3) | 3 (10.3) | 16.6 (12.9) |

Occupational Status
|  |  | Anxiety |  |  |  |  | Depression |  |  |  |  | Stress |  |  |  |  |
| --- | --- | --- | --- | --- | --- | --- | --- | --- | --- | --- | --- | --- | --- | --- | --- | --- |
|  |  | Non-Significant (0-10) | Mild (11-20) | Moderate (21-30) | Severe (31-40) | Mean (SD) | Non-Significant (0-7) | Mild (8-14) | Moderate (15-21) | Severe (22-28) | Mean (SD) | Non-Significant (0-10) | Mild (11-20) | Moderate (21-30) | Severe (31-40) | Mean (SD) |
| Student | 45 (46.8) | 18 (40.0) | 14 (31.1) | 8 (17.8) | 5 (11.1) | 26.7 (18.5) | 10 (22.2) | 15 (33.3) | 12 (26.7) | 8 (17.8) | 19.7 (14.9) | 14 (31.1) | 13 (28.9) | 12 (26.7) | 6 (13.3) | 22.3 (17.2) (18.5) |
| Employed | 62 (64.6) | 27 (43.6) | 21 (33.9) | 8 (12.9) | 6 (9.7) | 18.4 (15.1) | 23 (37.1) | 15 (24.2) | 16 (25.8) | 8 (12.9) | 13.67 (13.5) | 21 (33.9) | 13 (21.0) | 19 (30.6) | 9 (14.5) | 18.7 (14.5) |
| Retired | 7 (7.3) | 3 (42.9) | 0 (0.0) | 3 (42.9) | 1 (14.3) | 10.8 (7.3) | 2 (28.6) | 2 (28.6) | 2 (28.6) | 1 (14.3) | 9.0 (4.9) | 1 (14.3) | 3 (42.9) | 1 (14.3) | 2 (28.6) | 18.38 (8.1) |

**Figure 1b.**
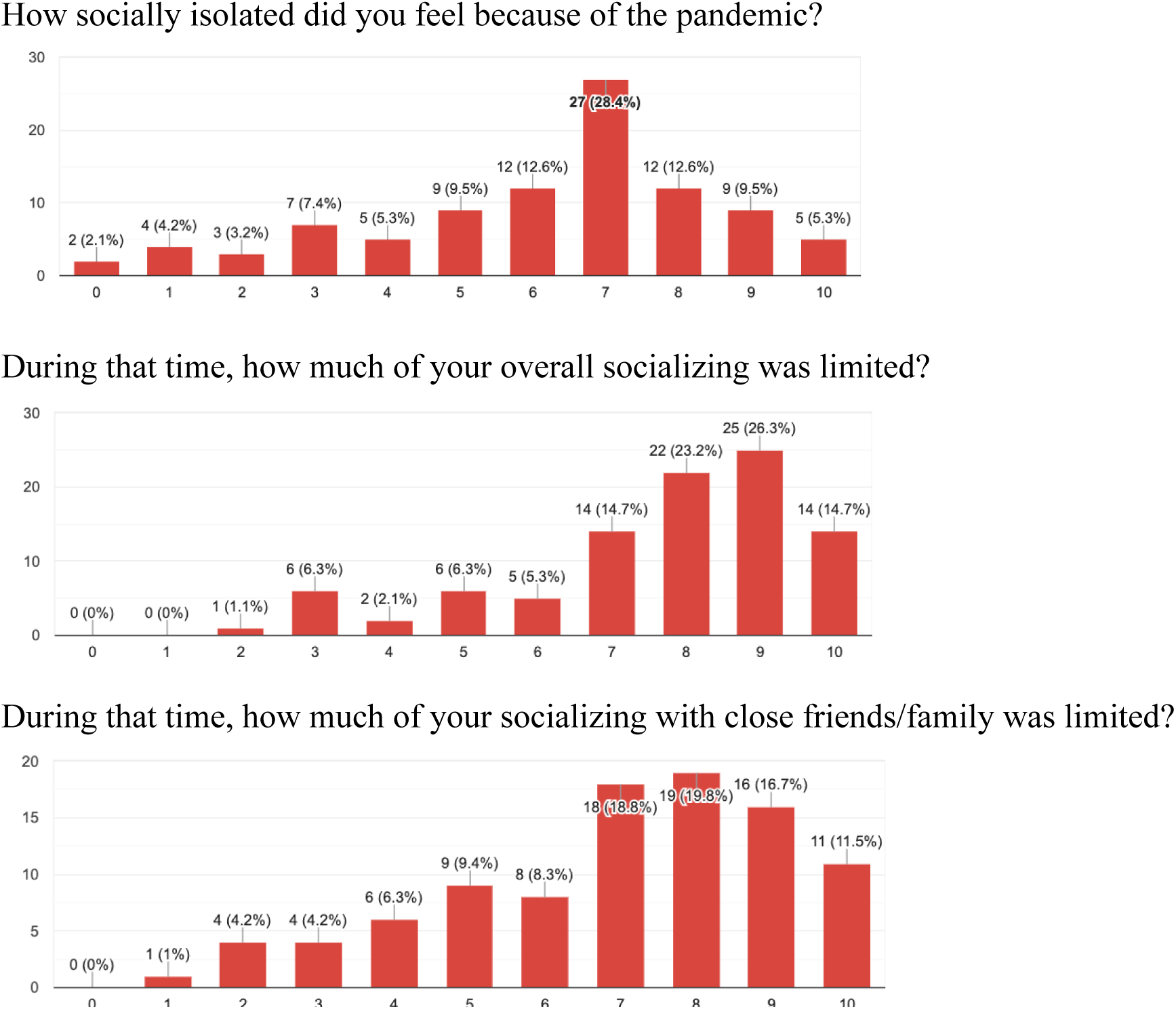
Responses on Social Isolation

**Figure 2b.**
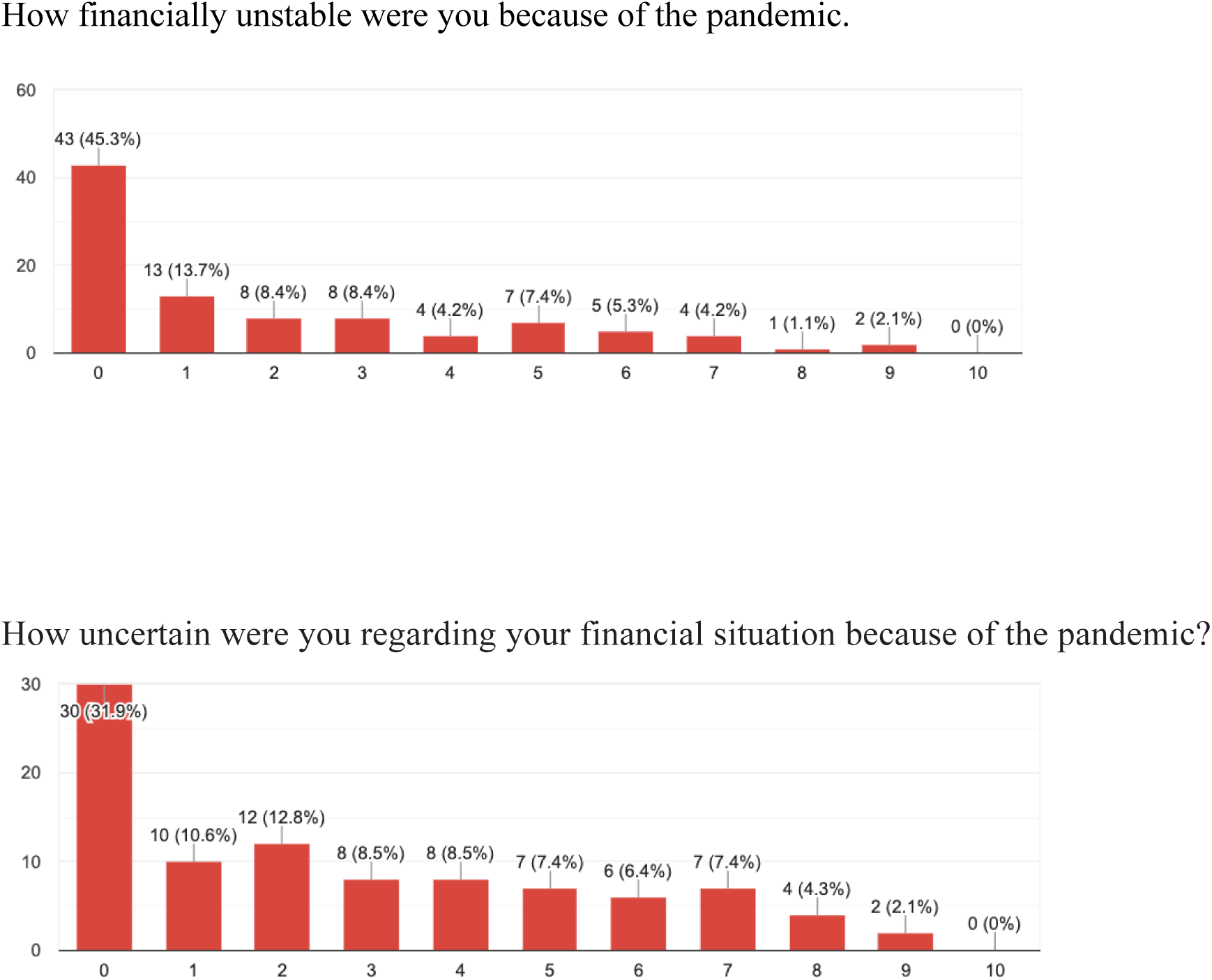
Responses on Financial Hardships

**Table 3b.** Responses on Health Concerns.

|  |  |
| --- | --- |
| I was infected with COVID-19 but easily recovered | 9 (9.4) |
| I became very ill due to COVID-19 | 0 (0.0) |
| I know someone personally who was infected with COVID-19 and easily recovered | 57 (59.4) |
| I know someone personally who became very ill from COVID-19 | 41 (42.7) |
| I know someone personally who died from COVID-19 | 24 (25) |
| There should be a complete lockdown of the country | 13 (13.5) |
| We should only rarely leave our houses, just for staples | 15 (15.6) |
| Social distancing and masks should be necessary, but there can still be a safe version of an open society | 66 (68.8) |
| I do not feel as though the general health of me and family/friends are in danger | 1 (1.0) |
| Feeling that too much unnecessary worry has been made about the COVID-19 outbreak | 5 (5.2) |

|  | Never | Less than half the time | Sometimes | More than half time | Always |
| --- | --- | --- | --- | --- | --- |
| Wearing a mask outside | 1 | 0 | 10 | 13 | 70 |
| Washing hands | 0 | 0 | 4 | 16 | 70 |
| Staying inside | 0 | 1 | 19 | 42 | 18 |

**Table 4b.** Multiple Linear Regression of Symptoms.

Anxiety Symptoms (GAD-7)
| | *B* | $\beta$ | 95% CI for *B* | p-value |
| --- | --- | --- | --- | --- |
| Gender (female, gender) | 1.12 | .094 | (0.82, 1.42) | .047 |
| Pre-existing chronic illness (yes vs. no) | 2.74 | .187 | (1.33, 4.07) | .04 |
| Complete quarantine (yes vs. no) | 1.56 | .103 | (.89, 2.23) | .223 |
| Infected with COVID-19 (yes vs. no) | -0.57 | -.089 | (-1.21, .47) | .001 |
| Leave house to work (yes vs. no) | -.19 | -.034 | (-1.84, 1.68) | .231 |
| Lost significant income (range from 0,1,2,3) | 2.9 | .173 | (-.07, 3.24) | .001 |

Depression Symptoms (PHQ-9)
| | *B* | $\beta$ |
| --- | --- | --- |
| Gender (female, gender) | 1.1 | .12 |
| Pre-existing chronic illness (yes vs. no) | 2.74 | .049 |
| Complete quarantine (yes vs. no) | 1.56 | .203 |
| Infected with COVID-19 (yes vs. no) | -.5 | .031 |
| Leave house to work (yes vs. no) | -.9 | -.3 |
| Lost significant income (range from 0,1,2,3) | 4.4 | .102 |

Traumatic Stress Symptoms (PSS)
| | *B* | $\beta$ |
| --- | --- | --- |
| Gender (female, gender) | 1.12 | .12 |
| Pre-existing chronic illness (yes vs. no) | 2.74 | .11 |
| Complete quarantine (yes vs. no) | 1.56 | .103 |
| Infected with COVID-19 (yes vs. no) | -.5 | -.08 |
| Leave house to work (yes vs. no) | -.6 | .19 |
| Lost significant income (range from 0,1,2,3) | 3.2 | .23 |

### Limitations

A significant degree of heterogeneity (in terms of scales, questionnaire as well as surrounding context regarding the level of pandemic when and where the studies were conducted) was noted throughout the studies assessed. Regarding the independent survey, a population size was smaller than expected and the study only included participants living in New York City which not only experienced a very severe pandemic and set of restrictions but represents one perspective compared to the meta-analysis which assessed 20 studies from 9 different countries, all of which experienced varying (quantitative/qualitative) degrees of restrictions and pandemic.

This study was performed in January 2020, 10 months into the COVID-19 shutdown. As with any pandemic, uncertainty is a driving factor. With the production and rollout of vaccinations, as of August 2021, more than 50% of the United States (where a significant number of studies utilized in this paper were performed, in addition to the generation population study) and one quarter of the world is fully vaccinated against SARS-Cov-2. With these statistics, the world, especially the United States, seemed to be turning a new corner of the pandemic, with fewer cases, hospitalizations, and deaths, masks mandates, social distancing and other manufactured facets of life seemed to be outdated and a sense of normalcy (regarding social life and health security) was returning to society. However, since July 2021 cases have begun to rise again due to a variant (Delta). This study is an accurate representation of prominent mental health symptoms from March 2020-January 2021. It should not and cannot be applied to current situations, even if certain aspects of the pandemic life (such as social restrictions and health concerns) overlap. The psyche is fluid and direct impacts/correlations are not well understood, and given the difference in situation and circumstances between different periods of time within the pandemic, assessments detailed above should not be considered comparable without thorough examination.

## Discussion

This meta-analysis study accompanied by a supplementary survey, investigated the levels of depression, anxiety and traumatic stress in the general population and found positive correlations between younger age and mental health symptoms, a greater impact on one’s mental health due to social isolation than health exposure, a significance of news exposure, a fluctuation in various issues of anxiety, depression and stress when assessing participants who had been infected, and a variation in time from change in lifestyle to expressed mental health symptoms depending on circumstances.

Depression symptoms between participants 18-25 years versus >60 years was 22.7% vs. 6.4%, a 238% increase (Ozamiz-Extebarria, 2020). Another study discovered significantly lower stress among the younger population (18-40) than participants older than 50 years (Qiu, 2020). People under 18 years had the lowest CPDI scores (a mean of 14.83 compared to the general population mean of 23.6 and the 27.8 mean for participants above 60 years). The five remaining studies revealed a consistent higher stress level in the younger population relative to the older population ranging from a 19% to a 267.6% increase. (Ahmed, 2020; Samardarshi, 2020; Smith, 2020; Shi, 2020; González-Sanguino, 2020; Wang, 2020). Older participants were less impacted by the lockdown order, as they are more likely to be retired and not as immersed in social activity. Yet, the increased health risk for the older population seems to be less impactive on anxiety and depression symptoms than the social isolation experienced by young adults.

Beyond the discrepancies of social isolation and health risk, an apparent difference between the young and old age groups is exposure to public information. Anxiety regarding health concerns was promoted by the consistent stream of information through social media and news. There was a clear correlation between an increased level of focus on the pandemic to increased prevalence of anxiety; a nearly 200% uptick in symptoms when participants consistently followed the pandemic on the news as opposed to never (Moghanibashi-Mansourieh, 2020; Huang, 2020). Participants who were less concerned about the media reports regarding the pandemic experienced 23.1% of the stress experienced by participants who were extremely concerned with the news and social media reports (Wang, 2020). Despite this increase in symptoms, symptoms nearly doubled for participants who felt as though they were not educated enough on the topic but felt as though their health was at risk. (Lei, 2020) Ironically, despite frontline workers experiencing a significant amount of anxiety and depression levels relative to the general population, their symptoms due to health concerns were negligent. This represents an apparent tug between regulated information and health concerns, and too little as well as too much knowledge is harmful. In addition, the fear of not knowing seemed to be more concerning and detrimental to one’s mental health than being too immersed in the issue. However, participants who were observed in the early stages on the pandemic when cases were increasing in other parts of the world, such as in February 2020 when the United States was seeing its first cases, lack of appreciation for the current state of the virus progression seemed to be beneficial for one’s mental health, which warrants the phrase “ignorance is bliss” (Olagoke, 2020).

The most novel and unexpected of findings was the inconsistent individual associations of anxiety, depression and traumatic stress experienced by participants previously infected with the COVID-19 virus. Participants who had been infected, compared to participants who had not, experienced decreased anxiety symptoms by 41.9%, increased depression symptoms by 107.7% and an increase in stress by 103.6%. Considering the hospitalization rate is only 14.8% and the death rate is 2.5%, most people infected with the disease survive without complications. The people who have suffered but survived the disease are more swayed by their realistic (low) risk of dying and not the mass health concerns which explains the decrease in anxiety symptoms. The increase in depression symptoms could be attributed to the heightened cognizance of the pandemic’s reality in contrast to the detached experience one would experience prior to infection.

During the first week of stay-at-home orders, the focus of 8 of the 20 studies revealed an increase of depression symptoms more than anxiety, but nearly all (86%) were due to disruptions of daily life, most significantly social interaction. The lockdowns were prompted by an alarming rise in COVID-19 cases (ranging from double to a ten-fold increase in weekly infections), yet the psychological response was centered on the impact of the safety measures (work at home, online school, reduced socialization) not the health cause. This became the facet of another correlation: the increase in age and reduction in symptoms. The more elderly participants (age >60), who were likely to be retired (7/10 pts.), did not experience a drastic change in lifestyle due to the shutdown; the transition to minimal social interaction would not be as impactful for those who already did not go to work. And despite the increased health risk, this absence of greater anxiety or distress thereformed confirmed the qualitative mental health impact centered on a reduction in social life.

The varying range of limited exposure witnessed in the studies consisted of a relationship between health risk and social isolation. Anxiety symptoms were nearly twice as prevalent when comparing participants who quarantined and did not (18.2% and 9.7%, respectively). However, when examining further, participants who had a direct occupational exposure risk were most vulnerable (27.2%) but participants who worked from home were more likely to experience symptoms of anxiety (15.2%) than participants who went to work with no direct contact with infected patients (7.9%) (Le Shi, 2020). The difference in lifestyle people who leave home for work are entitled to provides a benefit which compensates the increased health concerns from being more exposed to the disease.

## Data Availability

All relevant data are within the manuscript and its Supporting Information files

## Footnotes

1 Symptoms of depression, anxiety or stress refers to a score on one of the multiple self-report scales which indicates severity of moderate to high

## References

1. Brooke. (2020, June 9). Psychological inflexibility and intolerance of uncertainty moderate the relationship between social isolation and mental health outcomes during COVID-19 Brooke M. Smith a,*. Journal of Contextual Behavioral Science. https://wmich.edu/sites/default/files/attachments/news/2020/11/Smith%2C%20Twohy%2C%20Smith%2C%202020.pdf

2. Bruno. (2020, December 25). The Italian COVID-19 Psychological Research Consortium (IT C19PRC): General Overview and Replication of the UK Study. MDPI. https://www.mdpi.com/2077-0383/10/1/52

3. Centre for Addiction and Mental Health and the CAMH Foundation (2019). Screenings and Assessment. https://www.camh.ca/en/professionals/treating-conditions-and-disorders/anxiety-disorders/anxiety---screening-and-assessment

4. González-Sanguino. (2020, May 13). Mental health consequences during the initial stage of the 2020 Coronavirus pandemic (COVID-19) in Spain. PubMed Central (PMC). https://www.ncbi.nlm.nih.gov/pmc/articles/PMC7219372/

5. Huang. (2020, June 1). Generalized anxiety disorder, depressive symptoms and sleep quality during COVID-19 outbreak in China: a web-based cross-sectional survey. PubMed Central (PMC). https://www.ncbi.nlm.nih.gov/pmc/articles/PMC7152913/

6. Hyland. (2020, July 27). Anxiety and depression in the Republic of Ireland during the COVID-19 pandemic. Acta Psychiatrica Scandinavica. https://onlinelibrary.wiley.com/doi/full/10.1111/acps.13219

7. Lei Lei. (2020, April 26). Comparison of Prevalence and Associated Factors of Anxiety and Depression Among People Affected by versus People Unaffected by Quarantine During the COVID-19 Epidemic in Southwestern China. PubMed Central (PMC). https://www.ncbi.nlm.nih.gov/pmc/articles/PMC7199435/

8. Mazza. (2020, May 2). A Nationwide Survey of Psychological Distress among Italian People during the COVID-19 Pandemic: Immediate Psychological Responses and Associated Factors. PubMed Central (PMC). https://www.ncbi.nlm.nih.gov/pmc/articles/PMC7246819/

9. McBride. (2020, April 13). Monitoring the psychological impact of the COVID-19 pandemic in the general population: an overview of the context, design and conduct of the COVID-19 Psychological Research Consortium (C19PRC) Study. PsyArXiv Preprints. 10.31234/osf.io/wxe2n

10. Md Zahir. (2020, April 14). Epidemic of COVID-19 in China and associated Psychological Problems. PubMed Central (PMC). https://www.ncbi.nlm.nih.gov/pmc/articles/PMC7194662/

11. Mental Health, Substance Use, and Suicidal Ideation During the COVID-19 Pandemic. (2020, June 24). CDC. https://www.cdc.gov/mmwr/volumes/69/wr/pdfs/mm6932-H.pdf?deliveryName=USCDC_921-DM35222

12. Moghanibashi-Mansourieh. (2020, April 18). Assessing the anxiety level of Iranian general population during COVID-19 outbreak. PubMed Central (PMC). https://www.ncbi.nlm.nih.gov/pmc/articles/PMC7165107/

13. Moher. (2009, June 5). Preferred reporting items for systematic reviews and meta-analyses: the PRISMA statement. The BMJ. https://www.bmj.com/content/339/bmj.b2535

14. Olagoke. (2020, May 16). Exposure to coronavirus news on mainstream media: The role of risk perceptions and depression. PubMed Central (PMC). https://www.ncbi.nlm.nih.gov/pmc/articles/PMC7267047/

15. Ozamiz-Etxebarria. (2020, April 30). Stress, anxiety, and depression levels in the initial stage of the COVID-19 outbreak in a population sample in the northern Spain. Cadernos de Saúde Pública. https://scielo.br/scielo.php?script=sci_arttext&pid=S0102-311X2020000405013&lng=en&nrm=iso&tlng=en

16. Özdin. (2020, May 8). Levels and predictors of anxiety, depression and health anxiety during COVID-19 pandemic in Turkish society: The importance of gender. PubMed Central (PMC). https://www.ncbi.nlm.nih.gov/pmc/articles/PMC7405629/

17. Panchal. (2020, August 21). The Implications of COVID-19 for Mental Health and Substance Use. KFF. https://www.kff.org/coronavirus-covid-19/issue-brief/the-implications-of-covid-19-for-mental-health-and-substance-use/

18. Qiu. (2020, March 6). A nationwide survey of psychological distress among Chinese people in the COVID-19 epidemic: implications and policy recommendations. PubMed Central (PMC). https://www.ncbi.nlm.nih.gov/pmc/articles/PMC7061893/

19. Samadarshi. (2020, June 21). An online survey of factors associated with self-perceived stress during the initial stage of the COVID-19 outbreak in Nepal. Ethiopian Journal of Health Development. https://www.ajol.info/index.php/ejhd/article/view/201315

20. Shevlin. (2020, October 10). Anxiety, depression, traumatic stress and COVID-19-related anxiety in the UK general population during the COVID-19 pandemic. PubMed Central (PMC). https://www.ncbi.nlm.nih.gov/pmc/articles/PMC7573460/

21. Shi. (2020, July 1). Prevalence of and Risk Factors Associated With Mental Health Symptoms Among the General Population in China During the Coronavirus Disease 2019 Pandemic. Jama Network. https://jamanetwork.com/journals/jamanetworkopen/article-abstract/2767771

22. Wang. (2020, March 6). Immediate Psychological Responses and Associated Factors during the Initial Stage of the 2019 Coronavirus Disease (COVID-19) Epidemic among the General Population in China. PubMed Central (PMC). https://www.ncbi.nlm.nih.gov/pmc/articles/PMC7084952/

23. Wang. (2020, May 14). The psychological distress and coping styles in the early stages of the 2019 coronavirus disease (COVID-19) epidemic in the general mainland Chinese population: A web-based survey. PubMed Central (PMC). https://www.ncbi.nlm.nih.gov/pmc/articles/PMC7224553/

24. Wells. (2016). Ottawa Hospital Research Institute. The Ottawa Hospital Research Institute. http://www.ohri.ca/programs/clinical_epidemiology/oxford.asp

25. Xiong. (2020, August 8). Impact of COVID-19 pandemic on mental health in the general population: A systematic review. PubMed Central (PMC). https://www.ncbi.nlm.nih.gov/pmc/articles/PMC7413844/#bib0009

